# Stimulant Use Interventions May Strengthen ‘Getting to Zero’ Initiatives in Illinois: Insights from a Modeling Study

**DOI:** 10.1101/2021.09.22.21263980

**Authors:** Francis Lee, Daniel Sheeler, Anna Hotton, Natascha Del Vecchio, Rey Flores, Kayo Fujimoto, Nina Harawa, John A. Schneider, Aditya S. Khanna

## Abstract

**Objective(s):** Young Black men who have sex with men (YBMSM) are a key population identified in the Illinois Getting to Zero (GTZ) initiative who have experienced disproportionate HIV incidence. Rising stimulant use has been determined to impede the effectiveness of ART and pre-exposure prophylaxis for suppressing HIV transmission in populations. This modeling study explores the impact of stimulant use on HIV incidence among YBMSM – given the limited development of dedicated or culturally appropriate interventions for this population – and assesses the impact of these interventions on downstream HIV transmission in the context of achieving GTZ goals.

**Methods:** A previously developed agent-based network model (ABNM), calibrated using data for YBMSM in Illinois, was extended to incorporate the impact of stimulant use (methamphetamines, crack/cocaine, and ecstasy) on sexual networks and engagement in HIV treatment and prevention continua. The model simulated the impact of a residential behavioral intervention (BI) for reducing stimulant dependency and an outpatient biomedical intervention (mirtazapine) for treating methamphetamine dependence on improved engagement in the HIV treatment and prevention continua. The downstream impact of these interventions on population-level HIV incidence was the primary intervention outcome.

**Results:** Baseline simulated annual HIV incidence in the ABNM was 6.9(95% CI: 6.83,7.04) per 100 person years (py) and 453 (95% CI: 445.9,461.2) new infections annually. A residential targeted to 25% of stimulant users yielded a 27.1% decline in the annual number of new infections. Initiating about 50% of methamphetamine users on mirtazapine reduced the overall HIV incidence by about 11%. A 25% increase in antiretroviral treatment (ART) and preexposure prophylaxis (PrEP) uptake in the non-stimulant using YBMSM population combined with a 25% uptake of BI for stimulant users produces an HIV incidence consistent with HIV elimination targets (about 200 infections/year) identified in the GTZ initiative.

**Conclusions:** Targeted behavioral and biomedical interventions to treat stimulant dependency are likely to provide additive benefits to expanding ART and PrEP uptake for everyone in accomplishing GTZ initiatives for HIV elimination.

## INTRODUCTION

Getting to Zero (GTZ) Illinois is a HIV elimination strategy being implemented by a combination of state and county public health departments, academic medical centers, and community health organizations. GTZ Illinois assessments found that the overall declines in HIV incidence have not been experienced equally by sub-populations; younger (18-34 years) Black gay, bisexual and other MSM (YBMSM) have experienced relatively stable incidence rates over recent years.^1,2^The scale-up of antiretroviral treatment (ART) and preexposure prophylaxis (PrEP) use among YBMSM is a centerpiece of this elimination initiative and will require multi-level and combination-based interventions to realize the initiative goals.

The scale-up of ART and PrEP among YBMSM, however, is constrained because of the many psychosocial and healthcare barriers faced by YBMSM.^3,4^Substance use is one such barrier, and has been associated with suboptimal ART adherence and missed PrEP doses among MSM.^5–11^The use of stimulants – such as methamphetamines, crack/cocaine, and club drugs (e.g. ecstasy) – in particular, has been found to be associated with behaviors that may increase the risk of HIV transmission,^12^ particularly condomless insertive and receptive anal sex.^13^Black MSM living with HIV and not using methamphetamines have been found to be less likely to miss clinical visits for ART care than those who have used methamphetamines.^14^Emerging evidence also suggests that Black MSM who use social networking sites are often younger and more likely to have used methamphetamines and cocaine in the past 12 months compared to those who do not use such sites.^15^

Given the impact that stimulant use addiction plays in disengagement from HIV care, understanding some of the advances in treatment options available for stimulant use will be crucial to achieving GTZ policy goals. Mirtazapine, in particular, has been shown in clinical trials to be an effective biomedical treatment for methamphetamine addiction.^16,17^While no FDA approved treatment exists for treating cocaine addiction, other interventions such as residential rehabilitation have found moderate success in treating stimulant use disorders (including methamphetamines and cocaine).^18–21^The success of these interventions have led to calls for integration of HIV care to maximize the public health impact of these interventions.^22,23^

The GTZ Illinois planning committee has explicitly identified addressing substance use as a key component of their policy planning efforts to reduce the number of incident HIV cases among Black MSM in Illinois to a “functional zero” incidence, currently defined as fewer than 200 new infections per year.^24^ Fewer studies, however, have examined the prevalence of stimulant use and its role in HIV transmission among Black MSM specifically.^25^Transmission models that include the impact of stimulant use and sexual networks on the ART and PrEP continua can provide useful guidance for policy planning. One of the major challenges of addressing the stimulant epidemic among YBMSM is the development of culturally appropriate interventions for stimulant use treatment in this population, increasingly identified as a gap in the scientific literature and current public health policies in addressing psychosocial and structural barriers faced by Black populations.^26–29^

This study extends an existing agent-based network model (ABNM),^30^ parameterized largely with data collected in Illinois, to address the impact of stimulant use on the ART and PrEP continua and downstream HIV incidence among YBMSM. Interventions that are designed to treat stimulant use dependency (such as residential rehabilitation and medication-assisted treatment for methamphetamine use through mirtazapine) are thus likely to improve engagement in the HIV treatment and continua and are simulated to project their impact on HIV incidence and inform next steps in the GTZ planning efforts in Illinois.

## METHODS

### Agent-Based Network Model (ABNM) Development

The ABNM described below combines sexual network structure with a number of processes that impact HIV transmission. The sexual network structure was modeled using exponential random graph models (ERGMs),^31^ a statistically robust approach to model complex network evolution over time, and implemented using the *statnet*^32^ suite of packages in the R programming language. The ABM components were developed with the C++-based Repast HPC ABM toolkit.^33,34^Parameters and computer code to reproduce results are available in a public GitHub repository.^35^

### Demographic, Network, Behavioral and Biological Data

The baseline model was parameterized with data sources that were representative of YBMSM in Illinois. Local data sources included cohort data on Chicago YBMSM from “uConnect”^36,37^and the Young Men’s Affiliation Project (YMAP)^38,39^; both studies recruited participants in Chicago from 2013-2016 using systematic sampling schemes. Additional data on YBMSM were obtained from the National HIV Behavioral Surveillance (NHBS) survey in the Chicago Metropolitan Statistical Area.^40^ Other local and national sources, described below, were included where representative data from Illinois were not available. All procedures and protocols were approved by relevant institutional review boards.

### Baseline Model

Baseline HIV transmission was simulated to capture existing epidemic features among younger adults (age 18 to 34 years), populated with 10,000 individuals at the start of the dynamic simulations, approximately consistent with the number of estimated YBMSM in Chicago. The substantive model components included arrivals, departures, dynamic sexual network structure, the temporal evolution of CD4 counts and HIV RNA (“viral load”), HIV testing and diagnosis, dynamics of ART and PrEP use, external HIV infections, and HIV transmission dynamics (see Section A.4 of the Appendix for further detail).

### Modeling Impacts of Stimulant Use

#### HIV Treatment and Prevention Continua

The model examined the impact of methamphetamines, crack/cocaine and ecstasy on HIV treatment and prevention continua. Population-based cohort data were used to estimate the usage rates of methamphetamines, crack/cocaine and ecstasy.^36–39^ The model was seeded with users of the three classes of substances in accordance with the estimated rates. Estimates of ART adherence among users of the three substances were also derived from the available cohort data. The PrEP continuum for stimulant users was modeled in terms of reduced initiation and retention relative to the general population, as estimated in the literature.^5,41,42^ The ART and PrEP parameters for stimulant users are presented in Appendix Section 4.7. The key model parameters are listed in Table 1.

**Table 1:** Parameters to Model HIV Transmission among young Black men who have sex with men (YBMSM), Illinois.

|  |  |  |
| --- | --- | --- |
| **Demography** |  |  |
| **Parameter** | **Estimate** | **Source** |
| Age range | [18 – 34) years | Defined population of interest |
| HIV prevalence among 18-year old persons at entry into the model | 1% | <sup>48</sup> |
| Departures from simulated population | All agents achieving age > 34 years exit the population.<br>At any time in the simulation, agents uninfected with HIV experience mortality rates estimated in accordance with age-specific mortality for Chicago.<br>Agents infected with HIV experience mortality rates determined by their CD4 counts. | <sup>49</sup> |
| **Stimulant Use** |  |  |
| Rates of stimulant use (% of all simulated agents) | Methamphetamines: 4.0%<br>Crack/cocaine: 9.2%<br>Club drugs (e.g. ecstasy): 17.4% | <sup>36,37</sup> |
| **Sexual Behavior** |  |  |
| Mean partnership | All agents | <sup>40</sup> |
| duration | - Main: 512 days. Casual: 160 days.<br>- No difference assumed between stimulant users and non-stimulant users. |  |
| Mean number of main partnerships of stimulant users (per person) on any given day | Non-stimulant users: 0.38<br>Stimulant users<br>- Methamphetamines: 0.59<br>- Crack/cocaine: 0.69<br>- Club drugs (e.g. ecstasy): 0.61 | 36,37 |
| Mean number of casual partnerships (per person) on any given day | Non-stimulant users: 0.46<br>Stimulant users<br>- Methamphetamines: 0.59<br>- Crack/cocaine: 0.69<br>- Club drugs (e.g. ecstasy): 0.61 | 36,37 |
| **Adherence to Antiretroviral Treatment (ART)** |  |  |
| Distribution of ART adherence | Non-stimulant users:<br>- Never adherent: 10%<br>- Sometimes adherent: 30%<br>- Usually adherent: 28%<br>- Always adherent: 32%<br>Stimulant users<sup>*,†</sup>:<br>- Methamphetamines: 42% decline in percent always adherent<br>- Crack/cocaine: 50% decline in percent always adherent<br>- Ecstasy: 39% decline in percent always adherent | 36,37<br>Additional details on ART adherence in the overall population and among stimulant users are in Appendix Sections 4.7 and 4.9 respectively). |
| **PrEP Use** |  |  |
| Mean % of HIV-negatives who are prescribed PrEP on any given day | Non-stimulant users: 13.7%<br>Stimulant users:<br>- Methamphetamines: 5.4%<br>- Crack/cocaine: 7.1%<br>- Ecstasy: 4.6% | 36,37 |
| Mean time that a PrEP user is retained on PrEP | Non-stimulant users: 1 year<br>Stimulant Users: 9 months | 50<br>and additional details in Appendix Section 4.8 |
| Adherence to PrEP among PrEP initiators | Non-stimulant users<br>- Suboptimal adherence (0-3 pills/week): 38.1%<br>- High adherence (4+ pills/week): 61.9%<br>Stimulant users:<br>- Suboptimal adherence (0-3 pills/week): 76%<br>- High adherence (4+ pills/week): 24% | 51,52<br>Parameters for the overall population and stimulant users are derived in Appendix Sections 4.8 and 4.10 respectively |
| Reduction in transmission associated with levels of PrEP adherence | Non-adherence: 0%; low: 31%; moderate: 81%; high: 95% | 52,53 |
| <sup>*</sup>Persons always adherent to ART experiencing imperfect adherence due to stimulant use are uniformly distributed across the three other categories (usually, sometimes, and never adherent).<br><sup>†</sup>Users of multiple substances experience the highest declines associated with the substances of use. |  |  |

#### Sexual Behavior

Stimulant users in the model, identified by indicator variables denoting methamphetamine, crack/cocaine and ecstasy use, were given a propensity to form partnerships cross-sectionally that was greater than that of a non-user. This increased propensity was estimated by computing the ratio of the number of partnerships in the past six months for users of each of the stimulants relative to the number of partnerships reported by the overall YBMSM population (Table 1).

### Model Calibration

Model simulations proceeded in daily time steps. The model was calibrated over a 30-year period, using published HIV incidence and prevalence estimates as targets for calibration. Given the stochasticity in the model, each counterfactual setting was simulated 30 times to quantify the uncertainty in each simulated model run.

### Interventions

HIV incidence was measured over a 10-year period, where engagement in the HIV treatment and prevention continua and the sexual behavior of stimulant users were simulated as described above (**Baseline Model**). Comparing this outcome to HIV incidence in a hypothetical counterfactual with no stimulant use allowed for estimation of the impact of stimulant use on downstream HIV incidence (**Figure 1**).

**Figure 1:**
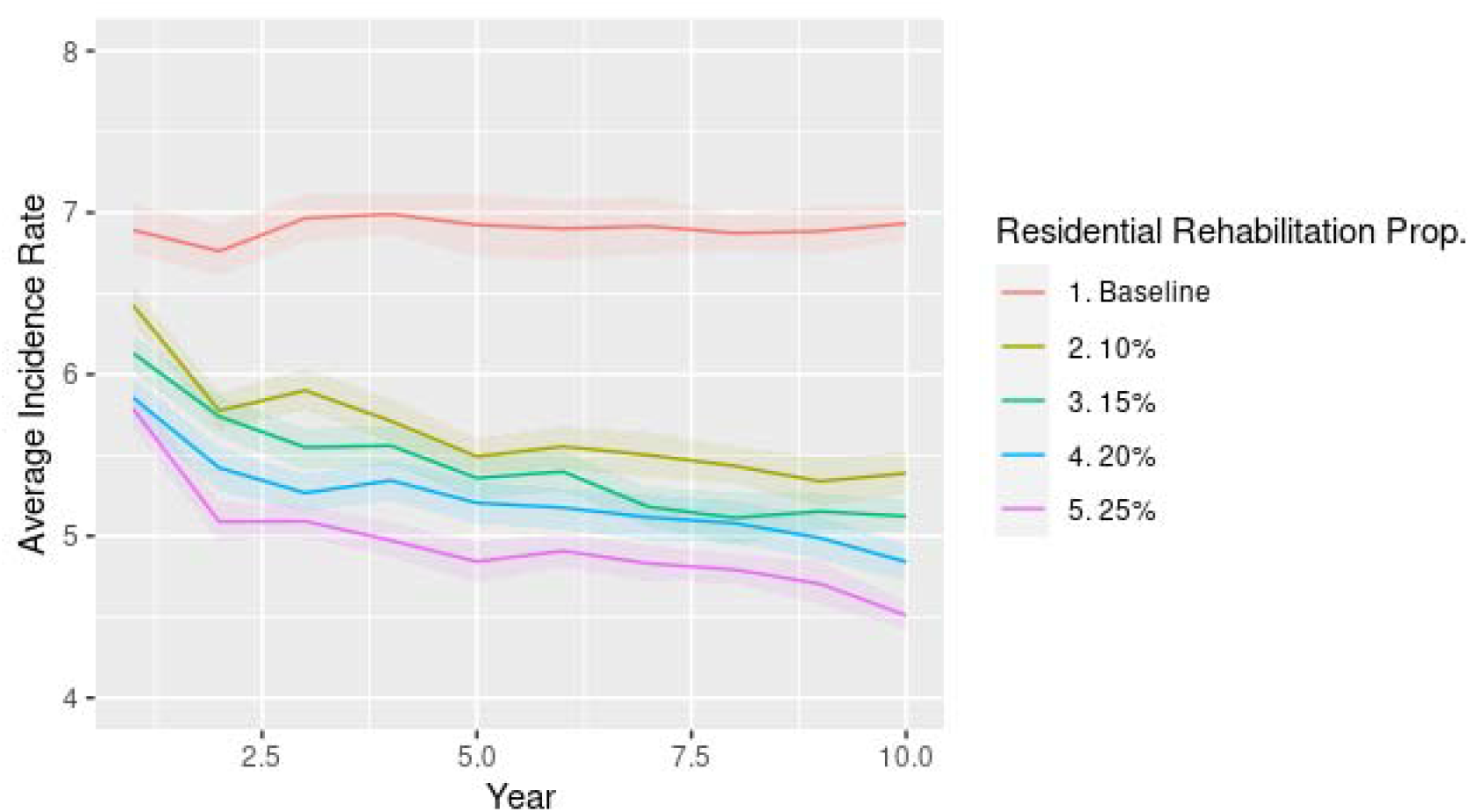
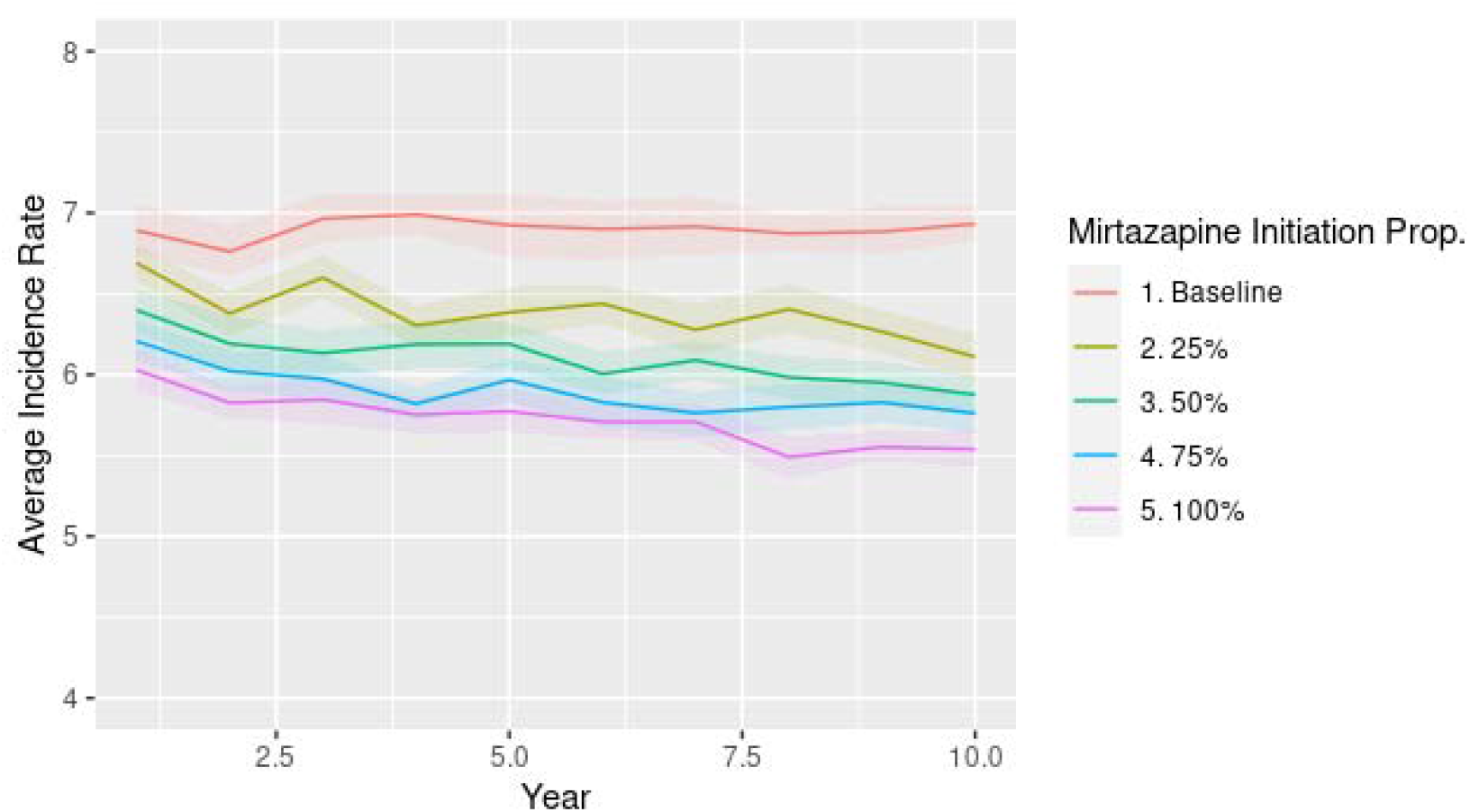
Mean population-level HIV incidence rate after implementation of:(top panel) the residential behavioral intervention (BI) for increasing proportions of stimulant users; (bottom panel) the mirtazapine intervention for increasing proportions of meth users.

A residential behavioral intervention (BI) for users of crack/cocaine, ecstasy and methamphetamine users was considered as an extension to the Baseline Model, similar to previous empirical studies that have demonstrated the impact of BIs on the sexual behavior of stimulant users.^43–45^ The impact of BI on stimulant dependency, which may improve downstream engagement in the HIV treat continua and its consequent impacts on population-level HIV incidence are less well-understood. In this study, we simulated an impact of a 3-month BI and its effects on engagement in the HIV treatment and prevention continua, consistent with typical durations of such interventions as implemented through a community rehabilitation program.^46^ HIV-undiagnosed persons who receive BI through a residential program are tested for HIV at the time of enrollment in the residential behavioral intervention.

Scenarios considering targeted BI for stimulant users are simulated, with the proportion of stimulant users receiving residential BI varied in separate counterfactuals at 10%, 15%, 20%, and 25%. (We limited the proportion of stimulant users receiving residential BI because it is likely to be an expensive intervention and wider scale-up may be limited by its cost). In accordance with empirical data, 87% of persons receiving residential BI benefit from it.^47^Thus, 87% of persons diagnosed with HIV who receive BI are assumed to be always adherent to ART during their period of residential stay because directly administered treatment and other structures support adherence in this setting (see Table 1 for the levels of ART adherence in the model). Similarly, 87% of HIV-negative persons receiving the BI intervention are assumed to be maximally adherent (4+ doses/week) to PrEP during the course of the residential BI intervention. Upon completion of BI, agents return to their pre-intervention levels of engagement in the HIV treatment and prevention continua.

Additionally, a biomedical intervention, consisting of mirtazapine for treating methamphetamine dependency, was simulated. During the period of biomedical treatment, 48.5% of the mirtazapine users were assumed to receive a mirtazapine outpatient prescription for a period of 3 months, consistent with common treatment mirtazapine treatment regimes.^16,17^ These persons who take mirtazapine as prescribed optimally adhere to their HIV medications (ART or PrEP) resulting in a 95% reduction in transmission of or risk for acquisition of HIV infection for the duration of their mirtazapine treatment (sensitivity analyses presented in Appendix Section A.6examine less than optimal engagement in the HIV treatment and prevention continua by the mirtazapine users). Those not adhering to mirtazapine as prescribed remained partially adherent (defined as an approximately 33% decline in infectivity or susceptibility) to their ART or PrEP prescriptions, respectively. Upon completion of the mirtazapine treatment, agents returned to their pre-treatment levels of methamphetamine use.

### Outcomes and Uncertainty Quantification

The primary outcomes for both residential BI and mirtazapine interventions were the mean number of HIV infections in the full population and the mean HIV incidence rate in the tenth year after the implementation at varying levels of uptake (Table 2). HIV incidence among methamphetamine users receiving mirtazapine was estimated for the various levels of uptake (Table 3). Additionally, scenarios that examined scale-up of ART and PrEP uptake in accordance with GTZ Illinois guidance, along with targeted stimulant use interventions, were modeled to assess the impact of stimulant use interventions on overall GTZ achievement targets (Table 4).

**Table 2:** Mean HIV incidence rate and the number of new HIV infections in the overall population in the tenth year after the implementation of the behavioral and biomedical interventions.

|  | 10<sup>th</sup> year HIV Incidence in the full population (per 100 person years) | New HIV Infections in 10th Year (full population) |
| --- | --- | --- |
| **Scenario** |  |  |
| *Baseline* | 6.93(6.83,7.04) | 453 (445,461) |
| *Behavioral Intervention (BI)* |  |  |
| Uptake<sup>*</sup> |  |  |
| 10% | 5.39 (5.26,5.52) | 379 (370,386) |
| 15% | 5.12 (5.02,5.23) | 365 (358,372) |
| 20% | 4.84 (4.72,4.95) | 348 (340,357) |
| 25% | 4.51 (4.42,4.6) | 330 (324,337) |
| *Mirtazapine* |  |  |
| Uptake<sup>**</sup> |  |  |
| 25% | 6.11 (5.96,6.25) | 411 (402,421) |
| 50% | 5.88 (5.74,5.99) | 402 (394,410) |
| 75% | 5.76 (5.64,5.88) | 400 (391,409) |
| 100% | 5.54 (5.43,5.66) | 387 (380,396) |
| <sup>*</sup>Proportion of stimulant users who receive the behavioral intervention (BI) |  |  |
| <sup>**</sup>Proportion of methamphetamine users who receive mirtazapine prescriptions |  |  |

**Table 3:** Mean HIV incidence rate and the number of new HIV infections among methamphetamine users in the tenth year after the implementation of the behavioral and biomedical interventions.

| Scenario | 10<sup>th</sup> year HIV Incidence among methamphetamine users (per 100) | New HIV Infections in 10th Year (methamphetamine users) |
| --- | --- | --- |
|  | person years) |  |
| *Baseline* | 10.63 (10.09,11.17) | 47 (45,49) |
| *Behavioral Intervention (BI)* |  |  |
| Uptake<sup>*</sup> |  |  |
| 10 | 7.14 (6.68,7.65) | 37 (35,40) |
| 15 | 6.34 (5.9,6.76) | 34 (32,37) |
| 20 | 5.92 (5.59,6.27) | 33 (31,35) |
| 25 | 5.58 (5.30,5.82) | 31 (30,33) |
| *Mirtazapine* |  |  |
| Uptake<sup>**</sup> |  |  |
| 25 | 7.18 (6.73,7.6) | 37 (35,39) |
| 50 | 6.37 (5.9,6.83) | 34 (31,37) |
| 75 | 5.66 (5.27,6.03) | 32 (30,35) |
| 100 | 4.71 (4.38,5.03) | 28 (26,30) |
| *Proportion of stimulant users who receive the Behavioral Intervention (BI) |  |  |
| **Proportion of methamphetamine users who receive mirtazapine prescriptions |  |  |

**Table 4:** Impacts of Behavioral Intervention (BI) for stimulant users on mean HIV incidence rate and the number of new HIV infections in the tenth year when ART and PrEP use are also scaled up for everyone.

|  |  |  |  |
| --- | --- | --- | --- |
| **ART and PrEP use for the full population** | **Targeted Behavioral Interventions for Stimulant Users<sup>\*</sup>** | **HIV Incidence in 10th Year (per 100 person years)** | **New HIV Infections in 10th Year** |
| 20% increase in ART and PrEP use across the full population over 10 years | None | 4.63 (4.52,4.75) | 329 (321,337) |
|  | 10% | 3.12 (3.04,3.21) | 240 (234,246) |
|  | 15% | 2.97 (2.9,3.03) | 230 (225,236) |
|  | 20% | 2.74 (2.67,2.82) | 215 (209,221) |
|  | 25% | 2.6 (2.52,2.68) | 205 (199,211) |
| 30% increase in ART and PrEP use across the full population over 10 years | None | 3.86 (3.77,3.94) | 281 (276,287) |
|  | 10% | 2.87 (2.78,2.97) | 222 (215,230) |
|  | 15% | 2.72 (2.66,2.78) | 212 (207,217) |
|  | 20% | 2.6 (2.52,2.68) | 205 (198,211) |
|  | 25% | 2.48 (2.39,2.57) | 197 (189,204) |
| *Proportion of stimulant users who receive BI |  |  |  |

Uncertainty in the HIV incidence projection estimates was quantified by using bootstrap estimates derived via simulation. To do this, the 30 simulation runs for each policy scenario at each time point were sampled 1,000 times with replacement. The mean for each of the resampled datasets was computed, and the 2.5% and 97.5% quantiles of these means were taken to obtain the 95% bootstrap confidence interval.

### Sensitivity Analyses

For residential BI, sensitivity analyses examined uncertainty in the proportion of stimulant users who receive the intervention, considering scenarios in which 10%, 15%, 20%, and 25% of stimulant users received BI. For mirtazapine, sensitivity analyses considered varying proportions of methamphetamine users receiving mirtazapine treatment. Scenarios where 5%, 25%, 50%, 75%, and 100% of methamphetamine users are given mirtazapine were simulated.

## RESULTS

Figure 1 provides the mean HIV incidence rate (per 100 person years) and the mean number of HIV infections in the ten years after the implementation of the BI intervention, with color bands that demonstrate the bootstrap confidence intervals. The control case, with no targeted intervention for stimulant users and ART and PrEP uptake maintained at baseline levels, yielded an average of 453.3 (95% CI: 445.9,461.2) new infections/year and a mean HIV incidence rate of 6.93 (95% CI: 6.83, 7.04) per 100 person years (py). In the tenth year, scaling up BI to 25% of stimulant users yielded a 27.1% decline in the number of new HIV infections to 330.5 (95%CI: 324.7, 337.3), and a 35.0% decline in the annual HIV incidence rate to 4.51 (95% CI: 4.42, 4.6) per 100 py (Table 2).

Figure 1 also shows the mean HIV incidence rate and the mean number of HIV infections in the full population ten years after the implementation of the mirtazapine intervention. The declines in overall HIV incidence are modest relative to the BI intervention: Providing mirtazapine to all meth users resulted in a 14.5% decline in the number of new HIV infections in the tenth year and a 20.1% decline in the HIV incidence rate (Table 2). This is not surprising because all stimulant users in the model (approximately 28% of the population) are eligible for BI, but a relatively smaller proportion (about 9% of the population) use methamphetamines and are therefore eligible for a mirtazapine prescription. In comparing the HIV incidence rate among methamphetamine users at various levels of uptake of BI and mirtazapine use, Table 3 shows that a 10% uptake of BI produces approximately the same HIV incidence as a 25% uptake of mirtazapine (about 7.2 per 100 person years). Similarly, comparable HIV incidence rates are produced among methamphetamine users when: (1) 15% of stimulant users receive the BI or 50% users of methamphetamines receive mirtazapine (about 6.3 per 100 py); and (2) 20% of stimulant users receive the BI or 75% users of methamphetamines receive mirtazapine (about 5.9 per 100 py and 5.7 per 100 py respectively).

Notably, stimulant use interventions are effective at achieving the functional zero HIV incidence target only when ART and PrEP is scaled up for everyone from the baseline uptake levels (Table 3). A 30% increase in ART and PrEP uptake in the general population combined with a 25% uptake of BI for stimulant users produces an HIV incidence of about 197 new infections per year. Additionally, a 30% increase in ART and PrEP uptake in the general population combined with a 20% uptake of BI for stimulant users, or a 20% increase in ART and PrEP uptake in the general population combined with a 25% uptake of BI for stimulant users, yields about 205 new infections per year, close to the target level of a functional zero HIV incidence.

## DISCUSSION

Our findings provide an assessment of the relative benefits of BI and mirtazapine for reducing HIV risk among people who use stimulants and the added benefits such policies can have in GTZ planning initiatives. On average, the residential BI implementation that at 10% uptake of all stimulant users (approximately 28% of the population) produced ewer new HIV infections annually than an outpatient mirtazapine intervention that reached all methamphetamine users (about 9% of the population). Approximately equal declines of HIV incidence among methamphetamine users are accomplished by a BI uptake of about 15% among users of any stimulants or a 50% uptake of mirtazapine among meth users.

Additionally, a 30% scale-up in ART and PrEP in the general population, combined with BI for stimulant users, has the potential to achieve a “functional zero HIV incidence” in 10 years. Previous modeling work has indicated that to get to a functional zero incidence of approximately 200 new HIV infections/year, ART and PrEP uptake in the overall population had to be scaled up by about 30% over 14 years.^24^ Here, we found that a targeted BI intervention of the type considered here may reduce that by four years.

Residential rehabilitation centers are likely to be expensive (approximately $215/day, according to some estimates^54^) and could be implemented via drug diversion programs or increased funding for voluntary addiction treatment. In addition to reducing negative health, social, and economic consequences of stimulant addiction, increased funding for addiction treatment could have cost benefits considering the high cost of HIV treatment.^55^Such analyses, however, are beyond the scope of this paper. Broader structural problems, such as food insecurity, housing instability, and mental illness comorbidities often impact the ability of stimulant users to engage in the ART and PrEP care continua. As residential drug rehabilitation facilities directly or indirectly address these problems while providing a structured environment, it is not surprising that engagement in the HIV treatment and prevention continua has been found to be higher during stay in a rehabilitation center. This study demonstrates that the effectiveness of a residential BI, even when persons undergoing treatment return to their baseline levels of engagement in the HIV treatment and prevention continua upon leaving the rehabilitation facility. In real life, the PrEP and ART engagement upon release from the facility is likely to be more varied; thus, the estimated effectiveness is likely to be a lower bound. Continued efforts to address the many barriers that impact long-term engagement in the HIV care and treatment continua are needed. Computational modeling can continue to provide much needed data on the implementation of interventions to address these barriers before they are implemented in Getting to Zero contexts.

We note several limitations in this study. First, interventions utilized in this model have been developed for White populations and more culturally appropriate interventions among YBMSM are needed.^26–29^We use a model that was developed for a YBMSM population, and future iterations of this work will consider the development and deployment of such interventions. Better contextual data on the association between stimulant use and engagement in the HIV prevention and treatment continua will be helpful in adapting these interventions for YBMSM. Second, this study modeled stimulant dependency as a binary variable. Future work might consider varying degrees of dependency among users of the stimulants that are considered here. Third, the financial costs of implementing behavioral and biomedical treatment programs for stimulant users, and the potential economic benefits of decarceration and rehabilitation were not examined here; such assessments will be important for future policymaking guidance. Fourth, future iterations of the model may consider contingency management and related interventions useful for treating stimulant dependency.^56–58^

This work begins to test empirically interventions in a simulation model designed for a YBMSM population. Future interventions, particularly culturally sensitive interventions for YBMSM who use stimulants are much needed, given their unique contexts of use, and the limited resources and safety nets for substance using YBMSM, relative to White populations. Achieving GTZ Illinois goal, for YBMSM may require addressing heterogeneities within YBMSM combined with a broad scale-up of biomedical prevention modalities and addressing the structural barriers that reduces the impact of such barriers. Direct treatment efforts to treat stimulant use, when implemented at scale, can help accomplish GTZ goals.

## Data Availability

All computer code and parameters needed to replicate the model are available in a public GitHub repository.

https://github.com/khanna7/BARS/tree/substance-use

## References

1. Illinois Department of Public Health. Data and Statistics: HIV Fact Sheets: Men Who Have Sex with Men.; 2016.

2. Illinois Department of Public Health. Data and Statistics: HIV Fact Sheet: Blacks.; 2014.

3. Hammett TM, Gaiter JL, Crawford C. Reaching seriously at-risk populations: health interventions in criminal justice settings. Health education & behavior□: the official publication of the Society for Public Health Education. 1998;25(1):99–120.

4. Bowleg L, Teti M, Malebranche DJ, Tschann JM. “It’s an Uphill Battle Everyday”: Intersectionality, Low-Income Black Heterosexual Men, and Implications for HIV Prevention Research and Interventions. Psychology of men & masculinity. 2013;14(1):25–34. doi:10.1037/a0028392

5. Hojilla JC, Vlahov D, Glidden D V, et al. Skating on thin ice: stimulant use and sub-optimal adherence to HIV pre-exposure prophylaxis. Journal of the International AIDS Society. 2018;21(3):e25103. doi:10.1002/jia2.25103

6. Grov C, Rendina HJ, John SA, Parsons JT. Determining the Roles that Club Drugs, Marijuana, and Heavy Drinking Play in PrEP Medication Adherence Among Gay and Bisexual Men: Implications for Treatment and Research. AIDS and behavior. Published online October 2018. doi:10.1007/s10461-018-2309-9

7. Wray TB, Chan PA, Kahler CW, Simpanen EM, Liu T, Mayer KH. Vulnerable Periods: Characterizing Patterns of Sexual Risk and Substance Use During Lapses in Adherence to HIV Pre-Exposure Prophylaxis among Men who have Sex with Men. Journal of acquired immune deficiency syndromes (1999). Published online November 2018:1. doi:10.1097/QAI.0000000000001914

8. Stimulant Use and Viral Suppression in the Era of Universal Antiretroviral Therapy. JAIDS Journal of Acquired Immune Deficiency Syndromes. 2019;80(1):89–93. doi:10.1097/QAI.0000000000001867

9. Carrico AW, Riley ED, Johnson MO, et al. Psychiatric risk factors for HIV disease progression: The role of inconsistent patterns of antiretroviral therapy utilization. Journal of Acquired Immune Deficiency Syndromes. 2011;56(2):146–150. doi:10.1097/QAI.0b013e318201df63

10. Hinkin CH, Barclay TR, Castellon SA, et al. Drug Use and Medication Adherence among HIV-1 Infected Individuals. AIDS and Behavior. 2007;11(2):185–194. doi:10.1007/s10461-006-9152-0

11. Marquez C, Mitchell SJ, Hare CB, John M, Klausner JD. Methamphetamine use, sexual activity, patient-provider communication, and medication adherence among HIV-infected patients in care, San Francisco 2004-2006. AIDS care. 2009;21(5):575–582. doi:10.1080/09540120802385579

12. Mimiaga MJ, Reisner SL, Fontaine Y-M, et al. Walking the line: stimulant use during sex and HIV risk behavior among Black urban MSM. Drug and alcohol dependence. 2010;110(1-2):30-37. doi:10.1016/j.drugalcdep.2010.01.017

13. Hermanstyne KA, Green HD, Tieu H-V, Hucks-Ortiz C, Wilton L, Shoptaw S. The Association Between Condomless Anal Sex and Social Support Among Black Men Who Have Sex With Men (MSM) in Six U.S. Cities: A Study Using Data from the HIV Prevention Trials Network BROTHERS Study (HPTN 061). AIDS and Behavior. 2018;1:3. doi:10.1007/s10461-018-2315-y

14. Hightow-Weidman L, LeGrand S, Choi SK, Egger J, Hurt CB, Muessig KE. Exploring the HIV continuum of care among young black MSM. Paraskevis D, ed. PLOS ONE. 2017;12(6):e0179688. doi:10.1371/journal.pone.0179688

15. Young SD, Shoptaw S. Stimulant Use Among African American and Latino MSM Social Networking Users. Journal of Addictive Diseases. 2013;32(1):39–45. doi:10.1080/10550887.2012.759859

16. Colfax GN, Santos GM, Das M, et al. Mirtazapine to reduce methamphetamine use: A randomized controlled trial. Archives of General Psychiatry. 2011;68(11):1168–1175. doi:10.1001/archgenpsychiatry.2011.124

17. Coffin PO, Santos GM, Hern J, et al. Effects of Mirtazapine for Methamphetamine Use Disorder among Cisgender Men and Transgender Women Who Have Sex with Men: A Placebo-Controlled Randomized Clinical Trial. JAMA Psychiatry. 2020;77(3):246–255. doi:10.1001/jamapsychiatry.2019.3655

18. AshaRani P, Hombali A, Seow E, Ong WJ, Tan JH, Subramaniam M. Non-pharmacological interventions for methamphetamine use disorder: a systematic review. Drug and Alcohol Dependence. 2020;212. doi:10.1016/j.drugalcdep.2020.108060

19. Gossop M, Marsden J, Stewart D. Treatment outcomes of stimulant misusers. Addictive Behaviors. 2000;25(4). doi:10.1016/S0306-4603(00)00063-0

20. Gossop M, Marsden J, Stewart D, Kidd T. Changes in use of crack cocaine after drug misuse treatment: 4–5 year follow-up results from the National Treatment Outcome Research Study (NTORS). Drug and Alcohol Dependence. 2002;66(1). doi:10.1016/S0376-8716(01)00178-8

21. Gossop M, Marsden J, Stewart D, Kidd T. The National Treatment Outcome Research Study (NTORS): 4-5 year follow-up results. Addiction. 2003;98(3). doi:10.1046/j.1360-0443.2003.00296.x

22. Sylla L, Bruce RD, Kamarulzaman A, Altice FL. Integration and co-location of HIV/AIDS, tuberculosis and drug treatment services. International Journal of Drug Policy. 2007;18(4). doi:10.1016/j.drugpo.2007.03.001

23. Farrell M, Martin NK, Stockings E, et al. Responding to global stimulant use: challenges and opportunities. The Lancet. 2019;394(10209). doi:10.1016/S0140-6736(19)32230-5

24. Khanna AS, Edali M, Ozik J, et al. Projecting the number of new HIV infections to formulate the “Getting to Zero” strategy in Illinois, USA. Mathematical Biosciences and Engineering. 2021;18(4):3922–3938. doi:10.3934/mbe.2021196

25. Okafor CN, Hucks-Ortiz C, Hightow-Weidman LB, et al. Brief Report: Associations Between Self-Reported Substance Use Behaviors and PrEP Acceptance and Adherence Among Black MSM in the HPTN 073 Study. JAIDS Journal of Acquired Immune Deficiency Syndromes. 2020;85(1). doi:10.1097/QAI.0000000000002407

26. Dangerfield DT, Cooper J, Heidari O, Allen S, Winder TJA, Lucas GM. Nursing and Health Care Preferences Among Opioid and Stimulant Using Black Sexual Minority Men: An Exploratory Study. Journal of the Association of Nurses in AIDS Care. 2021;32(5). doi:10.1097/JNC.0000000000000201

27. Sanchez K, Greer TL, Walker R, Carmody T, Rethorst CD, Trivedi MH. Racial and ethnic differences in treatment outcomes among adults with stimulant use disorders after a dosed exercise intervention. Journal of Ethnicity in Substance Abuse. 2017;16(4). doi:10.1080/15332640.2017.1317310

28. Sanchez K, Ybarra R, Chapa T, Martinez ON. Eliminating Behavioral Health Disparities and Improving Outcomes for Racial and Ethnic Minority Populations. Psychiatric Services. 2016;67(1). doi:10.1176/appi.ps.201400581

29. Saloner B, Cook BL. Blacks And Hispanics Are Less Likely Than Whites To Complete Addiction Treatment, Largely Due To Socioeconomic Factors. Health Affairs. 2013;32(1). doi:10.1377/hlthaff.2011.0983

30. Khanna AS, Schneider JA, Collier N, et al. A modeling framework to inform preexposure prophylaxis initiation and retention scale-up in the context of “Getting to Zero” initiatives. AIDS (London, England). 2019;33(12):1911–1922. doi:10.1097/QAD.0000000000002290

31. Recent developments in exponential random graph (p*) models for social networks. Social Networks. 2007;29(2):192–215. doi:10.1016/J.SOCNET.2006.08.003

32. Handcock MS, Hunter DR, Butts CT, Goodreau SM, Morris M. statnet: Software tools for the Statistical Modeling of Network Data. Published online 2003. http://statnetproject.org

33. Collier N, North M. Parallel agent-based simulation with Repast for High Performance Computing. Simulation. 2012;89(10):1215–1235.

34. Collier N, Murphy JT, Ozik J, Tatara E. Repast for High Performance Computing. Published online 2018. https://repast.github.io/repast_hpc.html

35. Khanna AS, Collier N, Ozik J. BARS: Building Agent-based Models for Racialized Justice Systems.

36. Preexposure prophylaxis awareness and use in a population-based sample of young black men who have sex with men. JAMA Internal Medicine. 2016;176(1). doi:10.1001/jamainternmed.2015.6536

37. Network dynamics of HIV risk and prevention in a population-based cohort of young Black men who have sex with men. Network Science. Published online February 2017:1–29. doi:10.1017/nws.2016.27

38. Kuhns LM, Hotton AL, Schneider J, Garofalo R, Fujimoto K. Use of Pre-exposure Prophylaxis (PrEP) in Young Men Who Have Sex with Men is Associated with Race, Sexual Risk Behavior and Peer Network Size. AIDS and Behavior. 2017;21(5):1376–1382. doi:10.1007/s10461-017-1739-0

39. Fujimoto K, Cao M, Kuhns LM, Li D, Schneider JA. Statistical adjustment of network degree in respondent-driven sampling estimators: Venue attendance as a proxy for network size among young MSM. Social Networks. 2018;54:118–131. doi:10.1016/j.socnet.2018.01.003

40. Paz-Bailey G, Mendoza MCB, Finlayson T, et al. Trends in condom use among MSM in the United States. AIDS. 2016;30(12):1985–1990. doi:10.1097/QAD.0000000000001139

41. Hojilla JC, Satre DD, Glidden D V., et al. Brief Report: Cocaine Use and Pre-exposure Prophylaxis: Adherence, Care Engagement, and Kidney Function. Journal of Acquired Immune Deficiency Syndromes. 2019;81(1):78–82. doi:10.1097/QAI.0000000000001972

42. Hojilla JC, Vlahov D, Crouch PC, Dawson-Rose C, Freeborn K, Carrico A. HIV Pre-exposure Prophylaxis (PrEP) Uptake and Retention Among Men Who Have Sex with Men in a Community-Based Sexual Health Clinic. AIDS and Behavior. 2018;22(4):1096–1099. doi:10.1007/s10461-017-2009-x

43. Wu E, El-Bassel N, Donald McVinney L, et al. Feasibility and Promise of a Couple-Based HIV/STI Preventive Intervention for Methamphetamine-Using, Black Men Who have Sex with Men. AIDS and Behavior. 2011;15(8):1745–1754. doi:10.1007/s10461-011-9997-8

44. Mimiaga MJ, Pantalone DW, Biello KB, et al. A randomized controlled efficacy trial of behavioral activation for concurrent stimulant use and sexual risk for HIV acquisition among MSM: project IMPACT study protocol. BMC Public Health. 2018;18(1):914. doi:10.1186/s12889-018-5856-0

45. Mimiaga MJ, Pantalone DW, Biello KB, et al. An initial randomized controlled trial of behavioral activation for treatment of concurrent crystal methamphetamine dependence and sexual risk for HIV acquisition among men who have sex with men. AIDS Care - Psychological and Socio-Medical Aspects of AIDS/HIV. 2019;31(9):1083–1095. doi:10.1080/09540121.2019.1595518

46. Moos RH, Moos BS, Andrassy JM. Outcomes of Four Treatment Approaches in Community Residential Programs for Patients With Substance Use Disorders. Psychiatric Services. 1999;50(12):1577–1583. doi:10.1176/ps.50.12.1577

47. Babudieri S, Dorrucci M, Boschini A, et al. Targeting Candidates for Directly Administered Highly Active Antiretroviral Therapy: Benefits Observed in HIV-Infected Injecting Drug Users in Residential Drug-Rehabilitation Facilities. AIDS Patient Care and STDs. 2011;25(6). doi:10.1089/apc.2010.0229

48. Waldo CR, McFarland W, Katz MH, MacKellar D, Valleroy LA. Very young gay and bisexual men are at risk for HIV infection: the San Francisco Bay Area Young Men’s Survey II. Journal of acquired immune deficiency syndromes (1999). 2000;24(2):168–174.

49. Centers for Disease Control and Prevention. CDC Wonder.

50. Preexposure Prophylaxis Initiation and Retention in Care Over 5 Years, 2012-2017: Are Quarterly Visits Too Much? Clinical infectious diseases. 2018;67(2):283–287. doi:10.1093/cid/ciy160

51. Liu AY, Cohen SE, Vittinghoff E, et al. Preexposure Prophylaxis for HIV Infection Integrated With Municipal- and Community-Based Sexual Health Services. JAMA internal medicine. 2016;176(1):75–84. doi:10.1001/jamainternmed.2015.4683

52. Incidence of Gonorrhea and Chlamydia Following HIV Preexposure Prophylaxis among Men Who Have Sex with Men: A Modeling Study. Clinical Infectious Diseases. Published online May 2017. doi:10.1093/cid/cix439

53. Grant RM, Anderson PL, McMahan V, et al. Uptake of pre-exposure prophylaxis, sexual practices, and HIV incidence in men and transgender women who have sex with men: a cohort study. The Lancet Infectious Diseases. 2014;14(9):820–829. doi:10.1016/S1473-3099(14)70847-3

54. Recovery Centers of America. Economic Cost of Substance Abuse in the United States, 2016. Retrieved September 10, 2021 from https://recoverycentersofamerica.com/economic-cost-substance-abuse/.

55. Metzger DS, Woody GE, O’Brien CP. Drug Treatment as HIV Prevention: A Research Update. JAIDS Journal of Acquired Immune Deficiency Syndromes. 2010;55(Supplement 1). doi:10.1097/QAI.0b013e3181f9c10b

56. Shoptaw S, Landovitz RJ, Reback CJ. Contingent Vs. Non-Contingent Rewards: Time-Based Intervention Response Patterns Among Stimulant-Using Men Who Have Sex With Men. Journal of Substance Abuse Treatment. 2017;72. doi:10.1016/j.jsat.2016.09.004

57. Corsi KF, Shoptaw S, Alishahi M, Booth RE. Interventions to Reduce Drug Use Among Methamphetamine Users at Risk for HIV. Current HIV/AIDS Reports. 2019;16(1). doi:10.1007/s11904-019-00423-y

58. Stitzer ML, Gukasyan N, Matheson T, et al. Enhancing patient navigation with contingent financial incentives for substance use abatement in persons with HIV and substance use. Psychology of Addictive Behaviors. 2020;34(1). doi:10.1037/adb0000504

