## Supplementary Appendix for "Stimulant Use Interventions May Strengthen ‘Getting to Zero’ Initiatives in Illinois: Insights from a Modeling Study"

### A.1. Introduction

This Supplementary Appendix describes additional technical components of the agent-based network model (ABNM) described in the main body of the manuscript, particularly focusing on providing a more detailed explanation of the model processes along with the parameters and data sources. Computer programs and supporting documentation are available at: <https://github.com/khanna7/BARS>.

The modeling methods utilized a structure similar to prior ABNMs of HIV transmission[1–5] following the following steps:

- An initial population was generated, as described in Section A.2 below.
- Main and casual partnership networks were simulated on this initial population. The modeling process is described in Section A.3.
- Baseline HIV epidemics, resulting from HIV infections transmitted through networks of main and casual partnerships, were simulated to capture features of the epidemic among young (18-34 years) black men who have with men (YBMSM). The steps mentioned in the main body of the paper to model the HIV epidemics are described in Section A.4.
- Diagnostics on our baseline model to ensure that it is well calibrated are presented in Section A.5.
- Sensitivity analysis for the mirtazapine intervention is presented in Section A.6.

### A.2 Initial Population

The initial population consisted of 10,000 individuals, uniformly distributed between the ages of 18-34 at the start, consistent with empirical data[6]. As time evolved, the rates of departure and entry of individuals were set so that the overall population grew slowly consistent with observed growth rates as per census data**.** The population was randomly seeded with 10% HIV prevalence at the start, sufficient to sustain an epidemic, as has been done in previous agent-ABNM studies to design HIV interventions [2,4,5,7]. The model was simulated over a long period (100 years) to allow epidemic outcomes to become consistent with empirical data; similar “burnin” periods have been instituted in previous ABNM studies [2,4,5,7]. The simulations incorporated a number of features pertaining to demography, sexual networks, biological, and behavioral features mentioned in the Methods section of the main body of the manuscript. More detail on the parameters describing these features is below.

### A.3 Models for Main and Casual Partnership Networks

The theoretical framework for modeling main and casual partnership networks is based upon the exponential random graph models (ERGMs), described elsewhere[8], and implemented in the *statnet* suite of packages [9] in the R programming language.

*Main and Casual Partnerships.* Two different partnership networks were simulated in the model. The log-odds of formation of each partnership type were dependent upon the number and distribution of existing partnerships within the network, and the absolute difference in the relative ages of partners for each partner type. The log-odds of the dissolution of each partnership were derived on estimates of the mean partnership duration.

The log odds of the formation of partnerships in both the main and casual networkswas specified as

$$\text{logit(p(}y_{ij,t}=1|Y_{ij,t-1}^{c},y_{ij,t-1}=0))=\theta\delta_{e}(e)+\sum_{d=0}^{2} \theta_{d}\delta_{d}\left( \eta\left( d \right) \right)\text{ + }\theta_{a}\delta_{a}\left( e\left| a_{i}-a_{j} \right| \right)+\sum_{k=1}^{3} \theta_{s_{k}}\delta_{s_{k}} N\left( s_{k} \right)\overline{d}\frac{\overline{d_{s_{k}}}}{d_{ns_{k}}}$$

where $i$ and $j$ are two individuals in the network; $t$ is a time step (simulated forward in daily units in this model), $t-1$ is the previous time step; $e$ is the number of edges; $\eta\left( d \right)$ is the number of nodes with degree $d$, specified for degrees 0, 1, and 2 in both the main and casual networks; and $\left( \left| a_{i}-a_{j} \right| \right)$is the difference in the absolute values of the ages of individuals who are in main and casual edges multipledby the number of main and casual partnerships$.$ The additional propensity of the substance users to form partnerships is calculating by multipling the mean number of partners per person for the full population $\overline{d}$multiplied by a scaling factor. This scaling factor considers the ratio of the estimated mean number of partners $\overline{d_{s_{k}}}$ for a specific substance $s_{k}$ to the mean number of partners for each person not using that substance $d_{ns_{k}}$. The propensity of partnership formation, as described above, is multiplied by the number of users of each of the three substances $N\left( s_{k} \right)$, to get the target number of partnerships for the users of the three substances.

The $\delta$ functions corresponding to each of these model terms represent the change in their value corresponding to the “toggle” of one dyad (defined as removing one existing tie, or adding a non-existent one); the change statistic functions are needed to estimate the coefficients $\theta$, $\theta_{d}\theta_{a}$ and $\theta_{s_{k}}$, corresponding to the edges, degree distribution age mixing terms, and propensity of substance users to form ties respectively. These coefficients are estimated using Markov Chain Monte Carlo techniques[10], as per the algorithmic routines contained in the *statnet* package [11].

For both main and casual networks, the persistence of each partnership was defined as

$$\text{l}\text{ogit(p(}y_{ij,t}=0|Y_{ij,t-1}^{c},y_{ij,t-1}=1))= \theta_{\mathrm{diss}}\delta_{e}(e)$$

where $\theta_{\mathrm{diss}}$is the coefficient associated with the dissolutoin of one tie. The model simulations, described in Section 4 below, incorporated the formation and dissolution coefficients derived here.

### A.4 Simulating Baseline Epidemics

The estimated models for main and casual partnership networks were simulated forward in daily time steps. These baseline epidemics were simulated for a long burnin period, allowing the model interdependencies sufficient time to equilibrate (100 years in this case). Each step of the simulation included the following processes: (1) entry of individuals into the modeled population; (2) departures of individuals from the modeled population;(3) modeling main and casual sexual networks; (4) HIV testing and diagnosis; (5) temporal evolution of CD4 counts; (6) temporal evolution of HIV RNA (“viral load”); (7) dynamics and effects of antiretroviral (ART) use; (8) dynamics and effects of preexposure prophylaxis (PrEP) use, (9) incidence of external HIV infections; and, (10) transmission of infection within serodiscordant main and casual partnerships. The estimation of the demographic, biological, and treatment parameters required to model these processes is described below.

A.4.1 Arrivals: Individuals aged into the model at 18 years. The arrival of new agents in the model was simulated as a Poisson process, with the mean set to 2.0. Thus, the entry rate was empirically determined, to balance the various departure processes (see Section A.4.2 below), so that the population grew at approximately the same growth rate as the population of interest (see Section A.4.2 below).

A.4.2 Departures and net population growth: Individuals departed from the model on account of the following reasons:

a. Aging out of the model at age >34 years;
b. HIV-uninfected individuals experience mortality, based on daily probabilities estimated from CDC Wonder data[12].
c. Untreated individuals with HIV infection had a maximum lifespan of 4279 days (approximately 11.7 years), estimated by summing the lengths of acute, chronic, and late-stage infection (details in Section A.4.10).
d. HIV-infected individuals on ART experienced an increase in the age-specific mortality rates, based on individual CD4 counts, in accordance with published data[13]. The increase in the daily mortality rates for HIV-infected individuals is below.

| **Table A.1. Increased mortality rates for HIV-infected individuals who are not using ART.** | |
| --- | --- |
| CD4 count (cells/µl) | %increase in age-specific mortality rates |
| < 50 | 51% |
| 50 – 99 | 37% |
| 100 – 199 | 26% |
| 200+ | 0% |

The arrival and departure processes described above resulted in a net simulated growth rate of 0.047%; FigureA.1 below shows the simulated population size over 10 years, averaged over 30 simulations, with PrEP initiation and retention held at baseline levels.

**Figure A.1: Simulated population size over 10 years, averaged over 30 simulations, with PrEP initiation and retention held at baseline levels.**

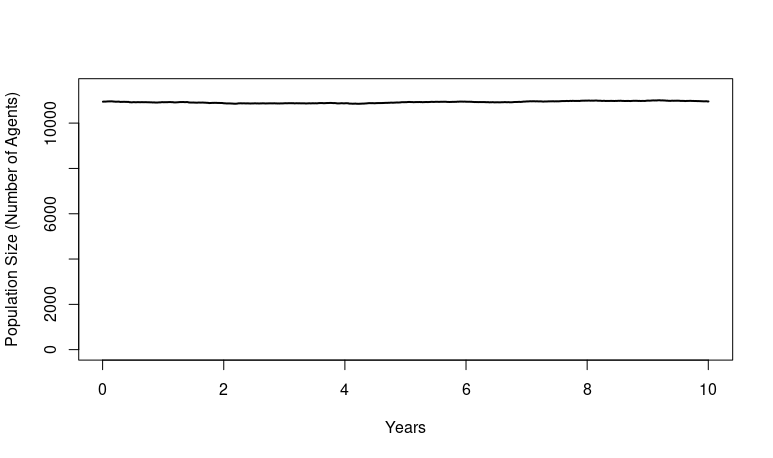

Since empirical data for the population growth rate of YBMSM specifically in Illinois were not available, we considered the population of young (18-35 years) Black males in Illinois, which grew at about 0.88% per year from 2010-2017, as per data provided by the U.S. Census Bureau [14]. Thus, both the modeled and empirical data revealed a population growth rate of <1% per year.

A.4.3 Sexual network structure: Two different sexual networks within this population were modeled, based on two types of partnerships: “main” and “casual”. The formation and dissolution processes of both partnership types were set such that their cross-sectional structure remained consistent with empirical data. The key parameters for main and casual partnerships, estimated for a given day, were: the mean number and distribution of partnerships, the mean partnership duration, and the mean of the absolute difference in ages for partners.

Sexual networks were dichotomized as “main” and “casual”. The underlying cohort data [15,16]used to parameterize these networks also contained information on “exchange” partners; these partnerships were modeled within the “casual” partnership typology. The same cohort data were used to estimate other parameters used to model the sexual networks, namely the mean number of cross-sectional main and casual partnerships, the distribution of the number of main and casual partnerships (0, 1, 2), and age-mixing. The degree distributions were estimated by computing overlaps in the dates of first and last sex between the study respondent and each of their partners (each respondent reported on up to 5 partners in the last six months). Because the age ranges of agents in our model spanned a relatively narrow range (18 – 34 years), the age-mixing parameter was estimated as the mean of the absolute values of the difference in the ages of the partners, in contrast to other models that were developed for broader age ranges and used the difference in the square-roots of partner ages [1,17]. The durations of mean and casual partnerships were estimated using the last partner reports from NHBS data [18].

A.4.4 Temporal evolution of CD4 counts: The CD4 count of uninfected men was assumed to be constant at 916 cells/µl [19].Upon HIV seroconversion, an individual’s CD4 count declined piecewise linearly, using a deterministic model where CD4 count was dependent on age at seroconversion, sex, and time since seroconversion, as described by Pantazis et al[20]:

$$\text{CD4}_{t}= \beta_{0}+\beta_{1}R+\beta_{2}F+t\left( \beta_{3}+\beta_{4}R+\beta_{5}A \right)^{2}$$

where $\text{CD}4_{t}$ was the CD4 count at $t$ years after seroconversion, $\beta_{0}$ was 23.53; $R$ was an indicator of African descent (set equal to 1 here), with a coefficient $\beta_{1}$ estimated at -0.76; $F$ was an indicator for female(set equal to 0here), $\beta_{3}$ estimated at -1.49 and $\beta_{4}$ estimated at 0.34; $A$ was the age at seroconversion, withcoefficient $\beta_{5}$ estimated at: 0 for $15\leq A<30$, -0.1 for $30\leq A< 34.$

A.4.5 Temporal evolution of HIV RNA (“viral load”): The viral load trajectory was modeled deterministically. For each infected, untreated individual, viral load was expressed as a six-parameter curve with a steep increase from 0 to peak viremia at 6.17 on the log_10_ scale over the first 45 days of infection, followed by a decline to the viral set point of 4.2 log_10_ over the next 45 days[21]. This viral set-point is maintained for the next 3550 days[22]. There is a final steady increase in viral load during late stage infection, where the viral load rises to 5.05 log_10_[23], over the course of 728 days[22]. The viral load of individuals who initiated ART decreased until treatment is interrupted, at which time the viral load increased again, as explained above, as long as treatment remained interrupted.

A.4.6 HIV testing and diagnosis: A heterogeneity of testing behaviors were modeled. Consistent with population-based cohort data, 7.8% of individuals <26 years and about 2.3% of individuals >=26 years of age were designated as never testing[24]. The remaining individuals were classified into categories defined by the frequency of testing. An individual was diagnosed if, at the time of testing, they had been infected for longer than the detection window of the test (i.e. 22 days[25]). The distribution of the number of HIV tests in the last two yearsis given in Table A.2below. Each individual was assigned to one of these categories, and a daily probability of testing for them was computed based on a number of tests parameter that was sampled from the discrete number of tests belonging to that interval.

| **Table A.2: Distribution of number of tests reported over 2 years** | |
| --- | --- |
| Number of tests in the last 2 years**^+^** | % of testers |
| 1-2 | 45.7% |
| 3-4 | 29.9% |
| 5-6 | 10.9% |
| 7-8 | 5.5% |
| 9-10 | 3.9% |
| 11-12 | 1.2% |
| 13-16 | 0.008% |
| 17-20 | 0.011% |
| 21-30 | 0.007% |
| ^+^Proportions reported based on Chicago YBMSM reporting ever testing for HIV. 7.8% of YBMSM younger than 25 and 2.3% of YBMSM older than 26 report never testing for HIV. | |

A.4.7 HIV Treatment Continuum: At the time of infection, each individual who was eligible for treatment was assigned to one of four states of ART adherence: adherence levels at the two extremes, almost always adherent (A), almost never adherent (N), and two categories of partial (P) adherence: usually(P+) and sometimes (P-). The distribution of ART adherence was estimated from longitudinal cohort data in the uConnect study [15,16]. Of the HIV-positive individuals completing all three visits (n=93), 32% were virally suppressed (<200 copies log viral load RNA) at all three visits (classified as category A defined above), 28% were suppressed at two visits (classified as category P+ above), 30% were suppressed at one visit (classified as category P- defined above), and 10% were never suppressed (classified as category N, defined above).

After 30 days, each person’s adherence for the next 30-day window was assessed, consistent with these four possible adherence states: 0.95 for A, 0.67 for P+, 0.33 for P-, and 0.05 for N. This cycle was repeated every 30 days given typical medication prescription patterns. (Typical medication prescription patterns for ART include a 30-day supply and 2-3 refills depending upon the client’s needs. This assumption was made in conjunction with our panel of HIV providers who care for YBMSM as well as the DHHS guidelines for ART treatment.)The distribution of times between diagnosis and ART initiation were empirically estimated from cohort data [15,16]and are given in Table 1 in the main body of the manuscript.

A.4.8 HIV Prevention Continuum: To model PrEP use, individuals were classified into four categories of adherence as per published data:21.1% of men took 0 pills/week (non-adherent), 7.0% took <2 pills/week (low adherence), 10.0% took 2–3 pills/week (moderate adherence), and 61.9% took 4+ pills/week (higher adherence)[26]. PrEP use is assumed to reduce HIV infection probability in these adherence groups by 0%, 31%, 81%, and 95%, for non, low, moderate, and high adherence, respectively, in accordance with previous modeling work [27]. On average, PrEP uptake (i.e., the proportion of HIV-negative individuals using PrEP at any given time) was about 13.7% [15,16]. To model this, consider probability $p$ of stopping PrEP on any given day. If $n$ is the number of HIV-negatives and $k$ is the proportion of HIV-negative individuals using PrEP, then on any given day $nkp$ is the number of users who stop PrEP. From the above definitions, it also follows that the number of HIV-negatives who are not using PrEP is $n\left( 1-k \right)$. If we define $q$as the probability that any non-user initiates PrEP on a given day,

$$n\left( 1-k \right)q = nkp$$

to maintain the same number of users at any given step, implying that $q =\frac{pk}{1-k}.$A selection probability of $q$ as defined above was set for HIV-negative individuals to initiate PrEP, enabling the model to maintain a specified proportion of HIV-negative individuals on PrEP.

For the interventions that prioritized serodiscordant couples and network position, the selection procedure for new PrEP initiators was implemented in two steps. In the first step, the process to maintain baseline PrEP rates was operationalized by assigning a selection probability $q$ for new PrEP initiators, as described above. In the second step, additional individuals were sampled from the pool of the target intervention group (serodiscordant couples or individuals in the highest scoring network positions) to make up the difference in the baseline and target levels of PrEP uptake. Selection from the target intervention group was also implemented probabilistically; the numerator of this probability was computed by considering the number of individuals required to make up the difference in the baseline and target levels of PrEP uptake, and the denominator was the total number of HIV-negative individuals.

Additionally, as per the base assumption in the model, PrEP initiators are retained on PrEP for 12 months, and retention times were assumed to be geometrically distributed. At the time of initiating PrEP use in the model, each user was assigned a duration of use, that was randomly sampled from a geometric distribution with a mean of 12 months. This formulation allowed PrEP users to cycle on and off PrEP, with the specified mean duration of use. Consequently, about 37% of PrEP initiators in the model were retained on PrEP one year after they started using it. A recent analysis of data from the largest provider of PrEP in Illinois found a comparable proportion (43% of PrEP initiators in the general population) retained on PrEP after about one year [28].

A.4.9 HIV Treatment Continuum for stimulant users: Analysis of population-based cohort data revealed that viral suppression rates among users of methamphetamines, crack/cocaine, and club drugs (such as ecstasy) were 42%, 50% and 39% respectively lower than the general population[15,16]. This decline in viral suppression rates was assumed to apply to the persons who are always adherent in the population. Thus, the proportion of HIV-diagnosed methamphetamines, crack/cocaine, and club drugs (such as ecstasy) users always adherent to ART was assumed to be 42%, 50% and 39% respectively lower than the general population. For users of multiple substances, the decline in ART adherence corresponded to whichever substance yielded the highest decline in ART adherence. For users of these stimulants, the decline in proportion of always adherent ART users was uniformly distributed across the other ART use categories: usually, sometimes, and never adherent. No change in the other elements of the ART continuum included in the model – namely, the HIV testing frequency and the lag time between HIV diagnosis and ART initiation – was assumed between the stimulant users and the general population.

A.4.10 HIV Prevention Continuum for stimulant users: For stimulant users, the PrEP continuum was modeled by estimating their mean adherence to and mean retention on PrEP. Our base model assumed that 61.9% of PrEP users were “optimally adherent”$\left( p_{a} \right)$, i.e., they took 4+ doses/week [29,30]. The remaining 38.1% of PrEP users, distributed across 0, 1, and 2-3 does/week, were “suboptimally adherent” $\left( 1- p_{a} \right)$. Prior research has suggested that stimulant users have a five-fold greater odds of suboptimal PrEP adherence than stimulant non-users [29]. Thus,

$$\frac{\frac{p_{a}}{1-p_{a}}}{\frac{p_{s}}{1-p_{s}}}=5$$

where $p_{s}$ denotes the proportion of stimulant users on PrEP are optimally adherent and $1-p_{s}$ denotes the proprtion of stimulant users on PrEP who are suboptimally adherent. Estimates from the cohort data above suggest that $\frac{p_{a}}{1-p_{a}}=1.56.$ Thus, the above equation can be solved to obtain $p_{s}=0.24$ and ${1-p}_{s}=0.76.$ Thus, about 76% of stimulant users on PrEP were estimated to be suboptimally adherent to PrEP.

A PrEP retention study has estimated that the ratio of the proportion of stimulant users who discontinue PrEP after one year to non-stimulant users who discontinue PrEP after 1 year is about 1.19 [30]. Our base model parameter was set to about 63% of PrEP users discontinuing PrEP within one year after initiation, comparable to estimates from clinic data in Chicago [28], as explained in Section A.4.8 above. Thus, about $1.19 \times63\%=75\%$of stimulant users who discontinue PrEP within one year of initiating it. Assuming that retention on PrEP is geometrically distribuetd, this estimate corresponds to a 9-month average retention on PrEP for stimulant users. For users of multiple substances, the decline in PrEP adherence and retention corresponded to whichever substance yielded the higher decline.

A.4.11.Incidence of external HIV Infections: To model HIV infections incident from non-YBMSM to the population (i.e. “external” infections), the following parameters were considered: (1) the overall incidence rate among YBMSM, estimated at 5-7 per 100 person years (py) from two different population-based Chicago cohorts of YBMSM[16,31,32]; (2) the proportion of this overall HIV incidence that consists of new HIV infections transmitted from older BMSM to YBMSM and vice versa, estimated at 28% [33]; the proportion of infections that are transmitted to YBMSM from older BMSM, assumed to be between 50% and 80%. Thus, the lower bound for a total number of infections incident externally among YBMSM in the model was 28%×50%×5 per 100 py = 0.70 per 100 py, and the upper bound was 28%×80%×7 = 1.60 per 100 py. The daily probability for each HIV-negative person to get externally infected thus ranged between $\frac{0.7}{365 \times100}$ and $\frac{1.6}{365 \times100}.$ Thus, this probability was used to conduct a Bernoulli trial for simulating an externally incident infection for each HIV-negative person at each time step. It was also assumed that the risk of getting externally infected was uniformly distributed with respect to age because this assumption produced simulated outcomes that were most consistent with empirical data.

Based on sensitivity analyses, we assumed that the risk of getting externally infected was uniformly distributed with respect to age because it produced the most consistent outcomes with the empirical data. External infections from women [34]and non-Black MSM [33]were not included due to evidence that very few infections among YBMSM are linked to either of these populations[34].

A.4.11. Transmission of HIV infection: Transmission of HIV infections through anal intercourse between HIV-infected and HIV-uninfected individuals within main and casual partnerships was modeled at each time step of the simulation. The probability of transmission during each sex act was derived using a number of different attributes, including stage of infection (determined by time since seroconversion) and viral load (in turn determined by the ART status of the infected partner) of the infected individuals, PrEP use by the uninfected partner, condom use at the time of intercourse (which reduced probability of transmission by 80%), and circumcision status of the uninfected partner.

Three stages of HIV infection were considered: acute, defined as lasting for 90 days from the onset of infection[21]; chronic infection that lasts for 3460 days [22]; and late-stage infection for 728 days [22]. Two widely varying estimates of the relationship between chronic stage infectivity and viral load were used in a sensitivity analysis (described in Section A.5 below) to determine the precise relationship between viral load and infectivity (defined as the probability of transmission during per act of condomless anal intercourse). The base infectivity during chronic HIV infection and additional multipliers for heightened infectivity during acute and late-stage infection are derived in Section A.5 below. The model included a 2.89-fold increase in infectivity corresponding to a unit increase in log viral load RNA[35].

### A.5 Model Diagnostics

Figure A.2 shows the prevalence and incidence trajectories of 30 stochastic model runs with the parameter estimates provided in Table 1 in the main body of the manuscript. To validate the simulation model, a number of outputs were	compared to empirically derived target values, as described in Table A.3 below.

**Figure A.2: Prevalence (A) and incidence (B) trajectories over thirty baseline model runs with the parameters listed in Table 1.***

(A) **Prevalence Trajectories**

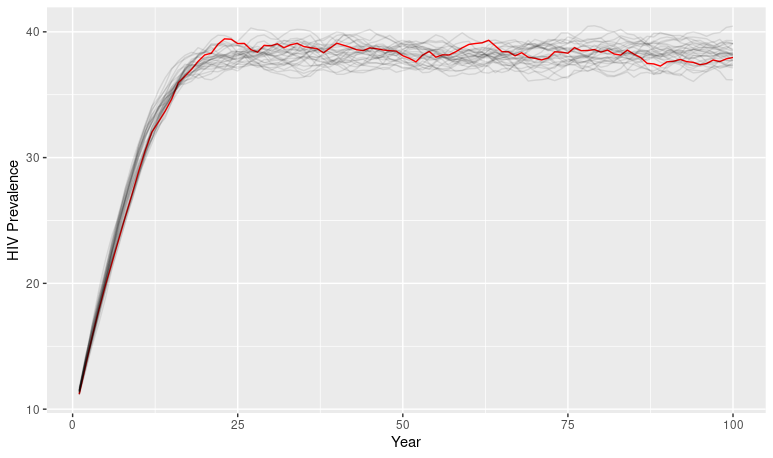

(B) **Incidence Trajectories**

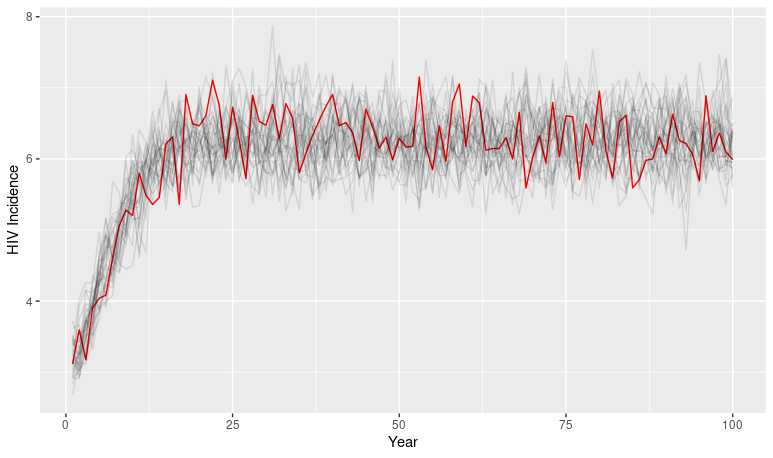

***The red curves denote the one instance that was selected for the intervention analyses.**

| **Table A.6: Comparison of simulated and target values for key model inputs for the full population.** | | | |
| --- | --- | --- | --- |
| **Parameter** | **Simulated statistics** | **Target statistics** | **References (for target statistics)** |
| Mean number of main partnerships per person | 0.41 | 0.38 | [15,16] |
| Mean number of casual partnerships per persons | 0.45 | 0.46 | [15,16] |
| Degree distribution for main partnerships | Degrees 0, 1, 2: 60%, 38.4%, 1.6% | Degrees 0, 1, 2: 64%, 34% 2% | [15,16] |
| Degree distribution for casual partnerships | Degrees 0, 1, 2: 61.6%, 31.6%, 6.7% | Degrees 0, 1, 2: 59%, 32%, 7% | [15,16] |
| Mean duration of main partnerships | 525.8 days | 512 days | [18] |
| Mean duration of casual partnerships | 163.8 days | 160 days | [18] |
| Per cent HIV-negatives on PrEP | 11.43% | 13.2% | [15,16] |

| **Table A.7: Comparison of simulated and target values for key model inputs for stimulant users.** | | | |
| --- | --- | --- | --- |
| **Parameter** | **Simulated statistics** | **Target statistics** | **References (for target statistics)** |
| Proportion of agents who use methamphetamine | 0.0903 | 0.092 | [15,16] |
| Proportion of agents who use crack/cocaine | 0.1765 | 0.1742 | [15,16] |
| Proportion of agents whose club drugs/ecstasy | 0.0452 | 0.0402 | [15,16] |
| Mean number of main and casual partnerships per person for meth users | Main:0.69  Casual: 0.72 | Main:0.69  Casual:0.71 | [15,16] |
| Mean number of main and casual partnerships per person for crack/cocaine users | Main: 0.58  Casual: 0.57 | Main:0.61  Casual:0.62 | [15,16] |
| Mean number of main and casual partnerships per person for club drug users | Main: 0.55  Casual: 0.59 | Main: 0.59  Casual:0.59 | [15,16] |
| PrEP initiation rates for stimulant users | Meth: 0.0556  Crack/Cocaine: 0.0506  Ecstasy 0.0448 | Meth: 0.071  Crack/Cocaine: 0.046  Ecstasy 0.054 | [15,16] |

### A.6 Sensitivity Analysis: Adherence to Mirtazapine Treatment

We perform a sensitivity analysis on the adherence to mirtazapine treatment among the methamphetamine users are prescribed. In accordance with empirical data that 48.5% of mirtazapine users took it as prescribed [38,39], a range of alternate adherence levels were examined, to consider less than optimal adherence scenarios: 39%, 29%, and 19% of mirtazapine users taking it as prescribed. The uptake levels of methamphetamine were considerd in accordance with the range of levels presented in the main body of the mansucript, i.e., scenarios with 0, 25%, 50%, 75%, and 100% of methamphetamine users were prescribed mirtazapine. The 10-year incidence for both the general population (Table A.8) and the methamphetamine using population (Table A.9) are presented below at each uptake proportion and adherence proportion.

| **Table A.8: HIV Incidence in the General Population**  **when Mirtazapine Adherence is Varied** | | | |
| --- | --- | --- | --- |
|  | Proportion taking Mirtazapine as Prescribed | | |
| Proportion of Methamphetamine Users Receiving Mirtazapine Prescriptions | 39% | 29% | 19% |
| 0% | 6.8 (6.7, 6.9) | 6.8 (6.6, 7.0) | 6.9 (6.7, 7.1) |
| 25% | 6.3 (6.1, 6.4) | 6.3 (6.2, 6.5) | 6.5 (6.3, 6.6) |
| 50% | 6.1 (5.9, 6.2) | 6.1 (6.1, 6.3) | 6.3 (6.1, 6.4) |
| 75% | 5.7 (5.6, 5.9) | 5.9 (5.8, 6.1) | 6.1 (6.0, 6.3) |
| 100% | 5.6 (5.4, 5.7) | 5.9 (5.7, 6.0) | 5.9 (5.7, 6.1) |

| **Table A.9: HIV Incidence among Methamphetamine Users**  **when Mirtazapine Adherence is Varied** | | | |
| --- | --- | --- | --- |
|  | Proportion taking Mirtazapine as Prescribed | | |
| Proportion of Methamphetamine Users Receiving Mirtazapine Prescriptions | 39% | 29% | 19% |
| 0% | 10.6 (10.0,11.2) | 10.8 (10.2,11.4) | 10.7 (10.0,11.4) |
| 25% | 7.3 (6.8,7.7) | 8.0 (7.5,8.5) | 8.0 (7.5,8.5) |
| 50% | 7.0 (6.5,7.5) | 7.0 (6.6,7.4) | 7.5 (7.0,7.9) |
| 75% | 6.3 (6.0,6.7) | 6.5 (6.1,6.9) | 6.9 (6.6,7.2) |
| 100% | 5.4 (5.1,5.7) | 6.1 (5.8,6.5) | 6.5 (6.0,6.9) |

### Relative to Table 3 in the main body of the manuscript, we see that declines in HIV incidence in the full population and among persons who use methamphetamines are dependent on the adherence to mirtazapine.

### A.7 References to the Appendix

1	Khanna AS, Goodreau SM, Gorbach PM, Daar E, Little SJ. **Modeling the Impact of Post-Diagnosis Behavior Change on HIV Prevalence in Southern California Men Who Have Sex with Men (MSM).** *AIDS and behavior* 2014; **18**:1523–31.

2	Khanna A, Goodreau SM, Wohlfeiler D, Daar E, Little S, Gorbach PM. **Individualized diagnosis interventions can add significant effectiveness in reducing human immunodeficiency virus incidence among men who have sex with men: insights from Southern California**. *Annals of Epidemiology* Published Online First: October 2014. doi:10.1016/j.annepidem.2014.09.012

3	Goodreau SM, Carnegie NB, Vittinghoff E, Lama JR, Sanchez J, Grinsztejn B, *et al.* **What Drives the US and Peruvian HIV Epidemics in Men Who Have Sex with Men (MSM)?** *PLoS ONE* 2012; **7**:e50522.

4	Khanna AS, Roberts ST, Cassels S, Ying R, John-Stewart G, Goodreau SM, *et al.* **Estimating PMTCT’s impact on heterosexual HIV transmission: A mathematical modeling analysis**. *PLoS ONE* 2015; **10**:e0134271.

5	Roberts ST, Khanna AS, Barnabas R V, Goodreau SM, Baeten JM, Celum C, *et al.* **Estimating the impact of universal antiretroviral therapy for HIV serodiscordant couples through home HIV testing: insights from mathematical models**. *Journal of the International AIDS Society* 2016; **19**. doi:10.7448/IAS.19.1.20864

6	United States Census Bureau. State Population by Characteristics: 2010-2016. 2017.

7	Khanna AS, Goodreau SM, Gorbach PM, Daar E, Little SJ. **Modeling the Impact of Post-Diagnosis Behavior Change on HIV Prevalence in Southern California Men Who Have Sex with Men (MSM)1 Khanna AS, Goodreau SM, Gorbach PM, Daar E, Little SJ. Modeling the Impact of Post-Diagnosis Behavior Change on HIV Prevalence** . *AIDS and behavior* 2014; **18**:1523–31.

8	Krivitsky PN, Handcock MS. **A Separable Model for Dynamic Networks**. *J R Stat Soc Series B Stat Methodol* 2014; **76**:29–46.

9	Handcock MS, Hunter DR, Butts CT, Goodreau SM, Morris M. statnet: Software tools for the Statistical Modeling of Network Data. 2003.http://statnetproject.org

10	Hunter DR, Handcock MS, Butts CT, Goodreau SM, Morris M. **ergm: A Package to Fit, Simulate and Diagnose Exponential-Family Models for Networks.** *Journal of statistical software* 2008; **24**:nihpa54860.

11	Goodreau SM, Handcock MS, Hunter DR, Butts CT, Morris M. **A statnet Tutorial.** *Journal of statistical software* 2008; **24**:1–27.

12	Centers for Disease Control and Prevention. CDC Wonder. 2017.http://wonder.cdc.gov/ucd-icd10.html (accessed 5 Dec2017).

13	May MT, Vehreschild J-J, Trickey A, Obel N, Reiss P, Bonnet F, *et al.* **Mortality According to CD4 Count at Start of Combination Antiretroviral Therapy Among HIV-infected Patients Followed for up to 15 Years After Start of Treatment: Collaborative Cohort Study.** *Clinical infectious diseases : an official publication of the Infectious Diseases Society of America* 2016; **62**:1571–1577.

14	United States Census Bureau. State Population by Characteristics: 2010-2017. 2018.https://www.census.gov/data/datasets/2017/demo/popest/state-detail.html (accessed 31 Jan2019).

15	Khanna AS, Michaels S, Skaathun B, Morgan E, Green K, Young L, *et al.* **Preexposure Prophylaxis Awareness and Use in a Population-Based Sample of Young Black Men Who Have Sex With Men.** *JAMA internal medicine* 2016; **176**:136–8.

16	Schneider J, Cornwell B, Jonas A, Lancki N, Behler R, Skaathun B, *et al.* **Network dynamics of HIV risk and prevention in a population-based cohort of young Black men who have sex with men**. *Network Science* 2017; :1–29.

17	Khanna A, Goodreau SM, Wohlfeiler D, Daar E, Little S, Gorbach PM. **Individualized diagnosis interventions can add significant effectiveness in reducing human immunodeficiency virus incidence among men who have sex with men: Insights from Southern California**. *Annals of Epidemiology* 2015; **25**. doi:10.1016/j.annepidem.2014.09.012

18	Paz-Bailey G, Mendoza MCB, Finlayson T, Wejnert C, Le B, Rose C, *et al.* **Trends in condom use among MSM in the United States**. *AIDS* 2016; **30**:1985–1990.

19	Witt MD, Lewis RJ, Rieg G, Seaberg EC, Rinaldo CR, Thio CL. **Predictors of the isolated hepatitis B core antibody pattern in HIV-infected and -uninfected men in the multicenter AIDS cohort study.** *Clinical infectious diseases : an official publication of the Infectious Diseases Society of America* 2013; **56**:606–12.

20	Pantazis N, Morrison C, Amornkul PN, Lewden C, Salata R a, Minga A, *et al.* **Differences in HIV natural history among African and non-African seroconverters in Europe and seroconverters in sub-Saharan Africa**. *PLoS ONE* 2012; **7**:e32369.

21	Little SJ, McLean AR, Spina CA, Richman DD, Havlir D v. **Viral dynamics of acute HIV-1 infection**. *Journal Of Experimental Medicine* 1999; **190**:841–850.

22	Buchbinder SP, Katz MH, Hessol NA, O’Malley PM, Holmberg SD. **Long-term HIV-1 infection without immunologic progression.** *AIDS (London, England)* 1994; **8**:1123–8.

23	Pilcher CD, Price MA, Hoffman IF, Galvin S, Martinson FEA, Kazembe PN, *et al.* **Frequent detection of acute primary HIV infection in men in Malawi**. *AIDS* 2004; **18**:517–524.

24	Khanna AS, Michaels S, Skaathun B, Morgan E, Green K, Young L, *et al.* **Preexposure Prophylaxis Awareness and Use in a Population-Based Sample of Young Black Men Who Have Sex With Men.** *JAMA internal medicine* 2016; **176**:136–8.

25	Branson BM, Stekler JD. **Detection of acute HIV infection: We can’t close the window**. *Journal of Infectioius Diseases* 2012; **205**:521–524.

26	Liu AY, Cohen SE, Vittinghoff E, Anderson PL, Doblecki-Lewis S, Bacon O, *et al.* **Preexposure Prophylaxis for HIV Infection Integrated With Municipal- and Community-Based Sexual Health Services.** *JAMA internal medicine* 2016; **176**:75–84.

27	Jenness SM, Goodreau SM, Rosenberg E, Beylerian EN, Hoover KW, Smith DK, *et al.* **Impact of the Centers for Disease Control’s HIV Preexposure Prophylaxis Guidelines for Men Who Have Sex With Men in the United States.** *The Journal of infectious diseases* 2016; **214**:1800–1807.

28	Rusie LK, Orengo C, Burrell D, Ramachandran A, Houlberg M, Keglovitz K, *et al.* **Preexposure Prophylaxis Initiation and Retention in Care Over 5 Years, 2012-2017: Are Quarterly Visits Too Much?** *Clinical infectious diseases* 2018; **67**:283–287.

29	Hojilla JC, Vlahov D, Glidden D v, Amico KR, Mehrotra M, Hance R, *et al.* **Skating on thin ice: stimulant use and sub-optimal adherence to HIV pre-exposure prophylaxis.** *Journal of the International AIDS Society* 2018; **21**:e25103.

30	Hojilla JC, Vlahov D, Crouch PC, Dawson-Rose C, Freeborn K, Carrico A. **HIV Pre-exposure Prophylaxis (PrEP) Uptake and Retention Among Men Who Have Sex with Men in a Community-Based Sexual Health Clinic**. *AIDS and Behavior* 2018; **22**:1096–1099.

31	Lancki N, Almirol E, Alon L, McNulty M, Schneider JA. **Preexposure prophylaxis guidelines have low sensitivity for identifying seroconverters in a sample of young Black MSM in Chicago.** *AIDS (London, England)* 2018; **32**:383–392.

32	Kuhns LM, Hotton AL, Schneider J, Garofalo R, Fujimoto K. **Use of Pre-exposure Prophylaxis (PrEP) in Young Men Who Have Sex with Men is Associated with Race, Sexual Risk Behavior and Peer Network Size**. *AIDS and Behavior* 2017; **21**:1376–1382.

33	Panneer N, Whiteside YO, France AM, Zhang T, Wertheim JO, Oster AM. Temporal Changes in HIV Transmission Patterns among Young Men Who Have Sex With Men, United States. Conference on Retroviruses and Opportunistic Infections (CROI). Session P-B1. 2017.

34	Oster AM, Wertheim JO, Hernandez AL, Cheryl M, Ocfemia B, Saduvala N, *et al.* **Using Molecular HIV Surveillance Data to Understand Transmission Between Subpopulations in the United States**. *J Acquir Immune Defic Syndr* 2015; **70**:444–451.

35	Hughes JP, Baeten JM, Lingappa JR, Magaret AS, Wald A, de Bruyn G, *et al.* **Determinants of per-coital-act HIV-1 infectivity among African HIV-1-serodiscordant couples**. *J Infect Dis* 2012; **205**:358–365.

36	Colfax GN, Santos GM, Das M, Santos DMD, Matheson T, Gasper J, *et al.* **Mirtazapine to reduce methamphetamine use: A randomized controlled trial**. *Archives of General Psychiatry* 2011; **68**:1168–1175.

37	Coffin PO, Santos GM, Hern J, Vittinghoff E, Walker JE, Matheson T, *et al.* **Effects of Mirtazapine for Methamphetamine Use Disorder among Cisgender Men and Transgender Women Who Have Sex with Men: A Placebo-Controlled Randomized Clinical Trial**. *JAMA Psychiatry* 2020; **77**:246–255.
